# Tall women with breast cancer have poorer survival than short women

**DOI:** 10.1101/2024.07.08.24310089

**Authors:** Steven Lehrer, Peter H. Rheinstein

**Affiliations:** Department of Radiation Oncology, Icahn School of Medicine at Mount Sinai, New York; Severn Health Solutions, Severna Park, Maryland

**Keywords:** breast cancer, height, genetics, risk, survival

## Abstract

**Background:** Tall women are more likely to develop breast cancer (BC). High Mobility Group AT-Hook 1(HMGA1), an oncofetal protein, plays a role in the progression of breast cancer. Non-coding sequences proximal to HMGA1 contain variants associated with 4.83 cm taller height. In the current study, we used UK Biobank data to examine the relationship of HMGA1 to height, risk, and prognosis of women with breast cancer.

**Methods:** Our analysis included all subjects with invasive BC that occurred either before or after participant enrollment and were recorded in the UK Biobank database using self-reported data and the International Classification of Diseases (ICD10, ICD9). We divided the subjects into three previously described three height groups: Short (< 155 cm), Medium (155 cm to 175 cm), Tall (> 175 cm). We analyzed the HMGA1 SNP rs41269028, a single nucleotide intron variant, C > T, minor allele frequency 0.044. SNP rs41269028 was previously evaluated in subjects with diabetes.

**Results:** Height of 9583 women with BC homozygous for the HMGA1 SNP rs41269028 major allele was 162.29 cm ± 6.18. Height of 944 women with BC who were carriers or homozygotes (CT + TT) of the minor allele T was 162.88 cm ± 6.001. This difference was significant (p = 0.005). The effect of height group on survival was significant (p = 0.032, log rank test). Tall women had the poorest survival. The effect of HMGA1 SNP rs41269028 genotype on BC risk (p = 0.602) and survival (p = 0.439, log rank test) was insignificant.

**Conclusion:** We conclude that HMGA1 influences height, but we were unable to demonstrate that HMGA1 is related to increased incidence or poor prognosis of tall women with breast cancer. We did find that tall women with breast cancer have poorer survival than short women. Our finding that tall women have a worse prognosis is important because it could help the oncologist decide, along with other prognostic factors, whether adjuvant therapy is warranted.

---

Tall women are more likely to develop breast cancer (BC) [1]. Women who are 176 cm or taller have a 20%–30% higher risk of breast cancer than women who are roughly 155 cm or lower, according to a pooled analysis that included data from 20 prospective cohort studies [2]. The growth spurts tall women experienced as children have been postulated to elevate risk of breast cancer associated with height. Increased hormone levels like IGF-1 or other growth factors can trigger growth spurts. Higher hormone levels and rapid cell proliferation during a growth spurt are thought to influence risk of breast cancer in later life.

High Mobility Group AT-Hook 1(HMGA1), an oncofetal protein, plays a role in the progression of breast cancer [3]. HMGA1 establishes an autocrine loop in invasive triple-negative breast cancer (TNBC) cells, which mediates the migration, invasion, and metastasis of TNBC cells and predicts the onset of metastasis in these patients.

Hawkes et al performed a whole genome sequencing association analysis for height using 333,100 individuals from three datasets: UK Biobank, TOPMed and All of Us. They identified non-coding sequences proximal to HMGA1 containing variants associated with a 4.83 cm taller height [4]. In the current study, we used UK Biobank data to examine the relationship of HMGA1 to height, risk, and prognosis of women with breast cancer.

## Methods

The UK Biobank is a large prospective observational study of men and women with no link to MedWatch. Participants were recruited from across 22 centers located throughout England, Wales, and Scotland between 2006 and 2010 and continue to be longitudinally followed for capture of subsequent health events [5]. This methodology is like that of the ongoing Framingham Heart Study [6], with the exception that the UKB program collects postmortem samples, which Framingham did not.

UK Biobank: has approval from the Northwest Multi-center Research Ethics Committee (MREC) to obtain and disseminate data and samples from the participants, and these ethical regulations cover the work in this study. Written informed consent was obtained from all participants. Details can be found at www.ukbiobank.ac.uk/ethics.

Our UK Biobank application was approved as UKB project 57245 (S.L., P.H.R.). Our analysis included all subjects with invasive BC that occurred either before or after participant enrollment and was recorded in the UK Biobank database using self-reported data and the International Classification of Diseases (ICD10, ICD9).

We divided the subjects into three previously described height groups [1]: Short (< 155 cm), Medium (155 cm to 175 cm), Tall (> 175 cm).

We analyzed the HMGA1 SNP rs41269028, a single nucleotide intron variant, C > T, minor allele frequency 0.044. SNP rs41269028 was previously evaluated in subjects with diabetes [7, 8].

Data processing was performed on Minerva, a Linux mainframe with Centos 7.6, at the Icahn School of Medicine at Mount Sinai. We used PLINK, a whole-genome association analysis toolset, to analyze the UKB chromosome files [9]. Statistical analysis was done with SPSS 26.

## Results

Data from 273,378 women, of which 10,527 were invasive breast cancer cases, was analyzed. Breast cancer patients were aged 60 ± 7 (mean ± SD). 95% of subjects were white British.

Table 1 shows HMGA1 SNP rs41269028 genotype versus height group in 8,327 post-menopausal breast cancer cases. A greater proportion of tall women with breast cancer (11.9%) than short women (6.8%) were carriers or homozygotes (CT + TT) of the minor allele T (p = 0.02, two tail Fisher exact test). No significant effect was present in pre-menopausal women (p = 0.441).

**Table 1.** HMGA1 SNP rs41269028 genotype versus height group in 8,327 post-menopausal breast cancer cases. A greater proportion of tall women with breast cancer (11.9%) than short women (6.8%) were carriers or homozygotes (CT + TT) of the minor allele T (p = 0.02, two tail Fisher exact test).

|  |  | CC | CT or TT |  |
| --- | --- | --- | --- | --- |
| short | Count | 1068 | 78 | 1146 |
|  | % within height group | 93.2% | 6.8% | 100% |
| medium | Count | 6414 | 632 | 7046 |
|  | % within height group | 91.0% | 9.0% | 100% |
| tall | Count | 119 | 16 | 135 |
|  | % within height group | 88.1% | 11.9% | 100% |
| total | Count | 7601 | 726 | 8327 |
|  | % within height group | 91.3% | 8.7% | 100% |

Height of 9583 women with BC homozygous for the HMGA1 SNP rs41269028 major allele (CC) was 162.29 cm ± 6.18. Height of 944 women with BC who were carriers or homozygotes (CT + TT) of the minor allele T was 162.88 cm ± 6.001. This difference was significant (p = 0.005).

Figure 1 illustrates 8,327 post-menopausal breast cancer patients stratified by height group and HMGA1 SNP rs41269028 genotype (CC versus CT or TT).

**Figure 1.**
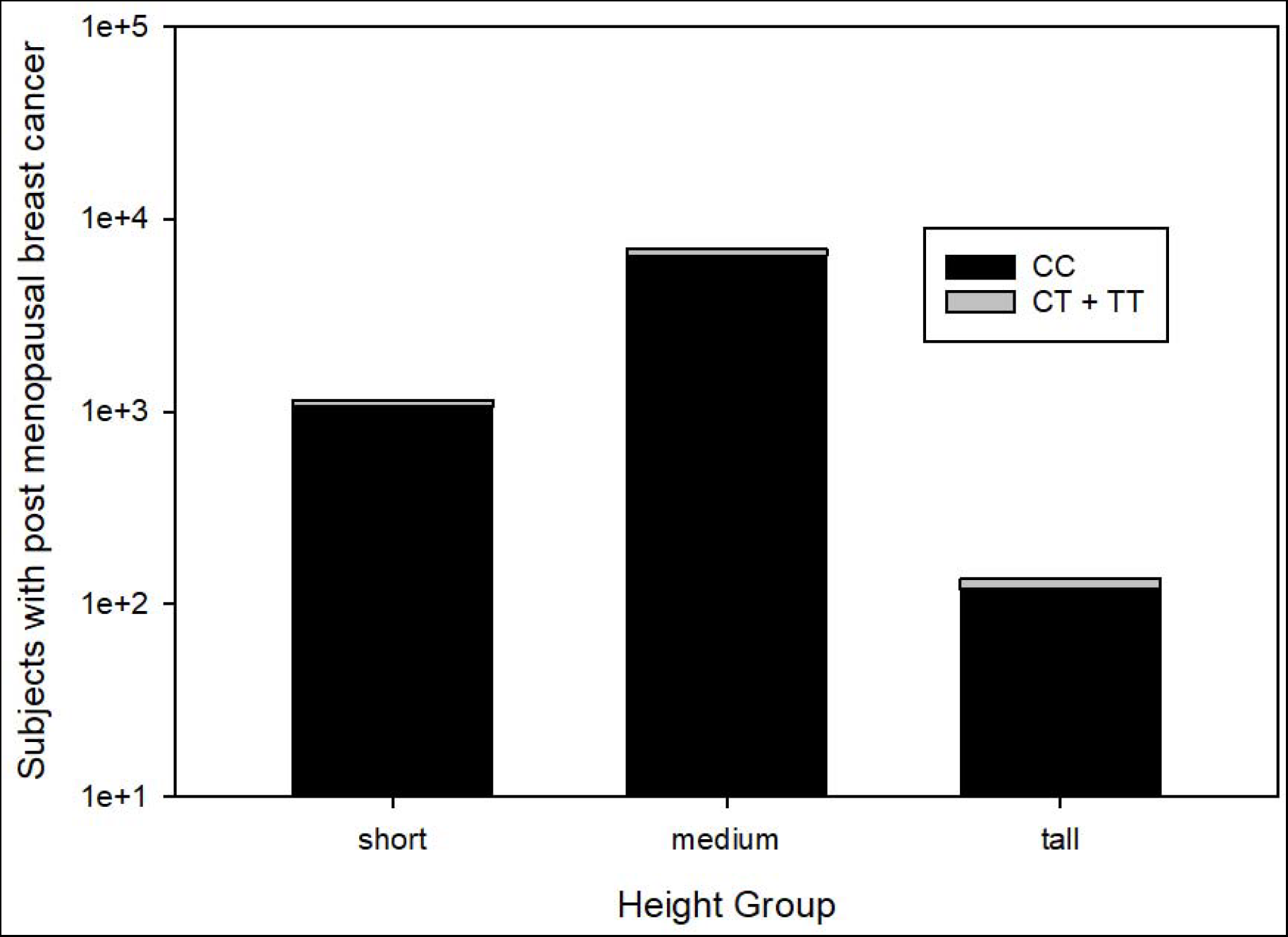
8,327 post-menopausal breast cancer cases stratified by height group and HMGA1 SNP rs41269028 genotype (CC versus CT or TT). Note that tall women have the greatest proportion of carriers or homozygotes (CT + TT) for the minor allele T (p = 0.02, two tail Fisher exact test).

Aging is associated with height loss [10]. To correct for this effect, multivariate linear regression was performed on breast cancer cases, height group dependent variable, HMGA1 SNP rs41269028 genotype and age independent variables. The effect of genotype on height groups was significant (B = 0.040, p = 0.002) and independent of the effect of age (B = - 0.005, p < 0.001). In other words, carriers or homozygotes of the minor allele T were taller than homozygotes for the major allele (CC); while older women were shorter than younger women.

Figure 2 illustrates survival of breast cancer subjects stratified by HMGA1 SNP rs41269028 genotype. The effect of genotype was insignificant (p = 0.439, log rank test).

**Figure 2.**
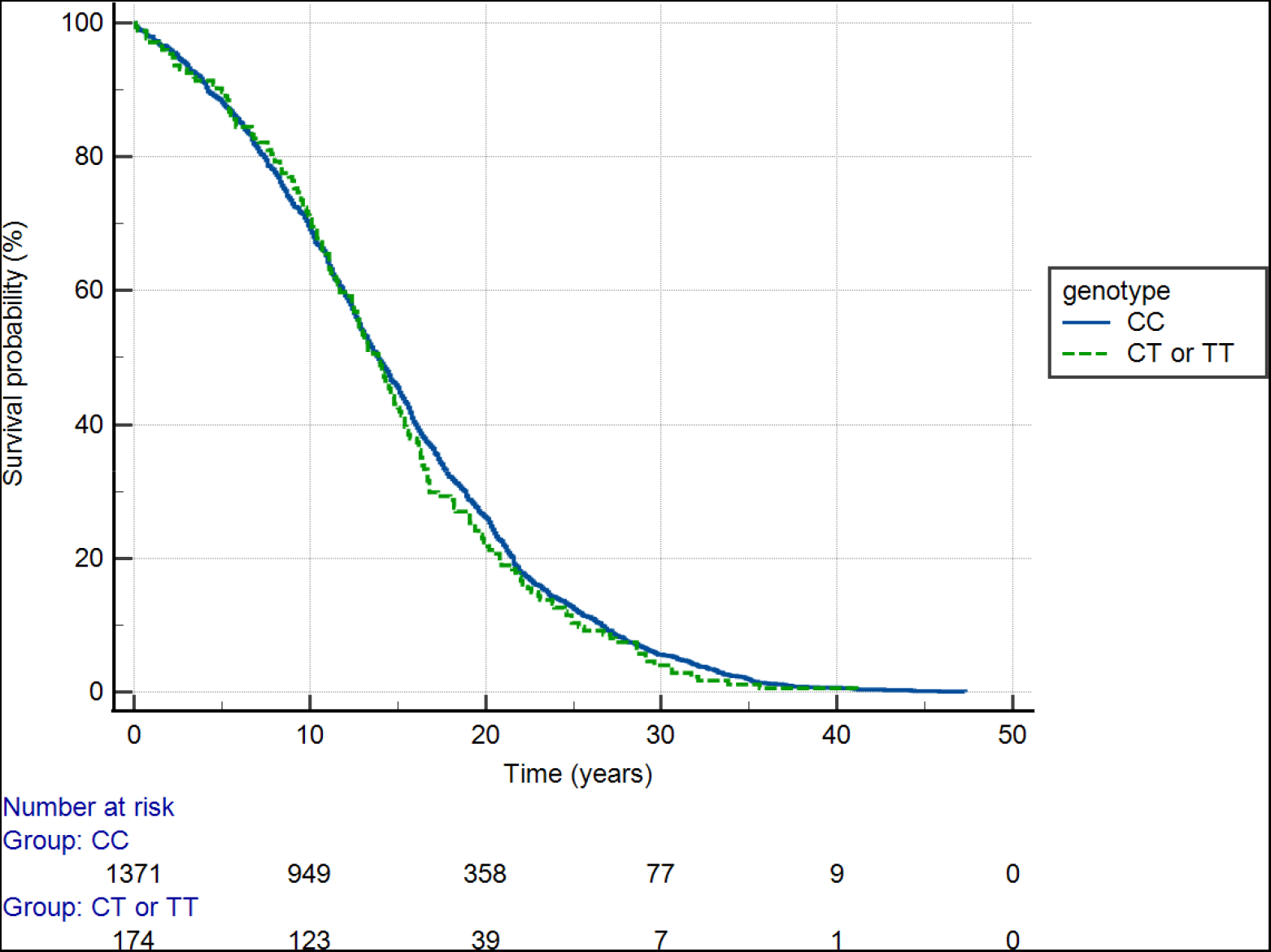
Survival of breast cancer subjects stratified by HMGA1 SNP rs41269028 genotype. The effect of genotype was insignificant (p = 0.439, log rank test).

Figure 3 shows survival of breast cancer subjects stratified by height group. The effect of height group was significant (p = 0.032, log rank test). Tall women had the poorest survival.

**Figure 3.**
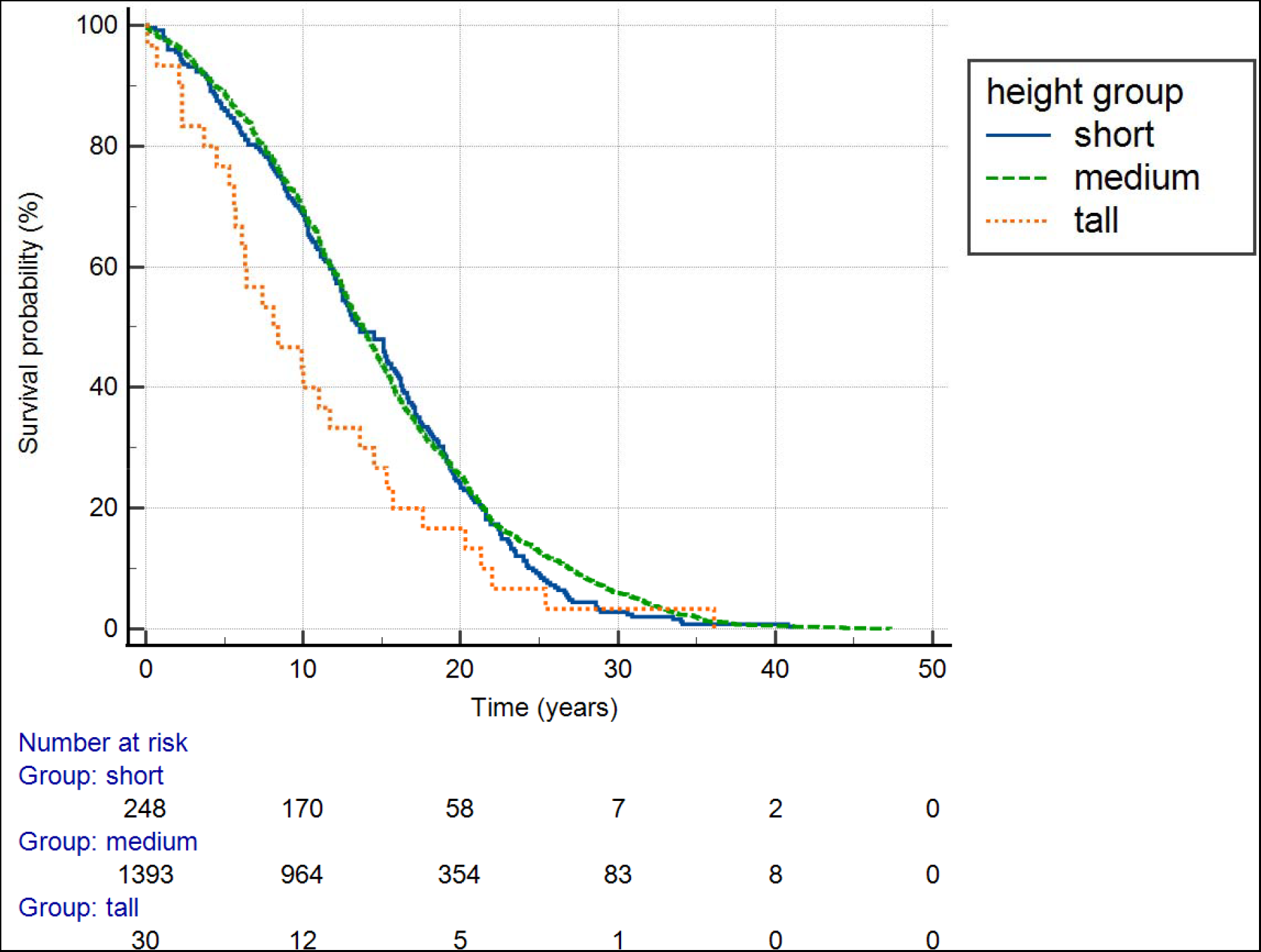
Survival of breast cancer subjects stratified by height group. The effect of height group was significant (p = 0.032, log rank test). Tall women had the poorest survival.

Table 2 has results of logistic regression, O.R. odds ratio, 228,611 women, breast cancer yes or no dependent variable, age, menopause status, height group, independent variables. Risk of breast cancer was increased in post-menopausal women (O.R. 4.315, p < 0.001). Risk increased with each year of age (O.R. 1.034, p < 0.001). Short women were at decreased risk compared to tall women (O.R. 0.819, p = 0.026). Women of medium height were at decreased risk that was not significant compared to tall women (O.R. 0.929, p = 0.391). HMGA1 SNP rs41269028 had no significant relationship to BC risk (p = 0.602).

**Table 2.** Logistic regression, L.B. lower bound, U.B. upper bound, O.R. odds ratio, 228,611 women, breast cancer yes or no dependent variable, age, menopause status, height group, independent variables. Risk of breast cancer was increased in post-menopausal women (O.R. 4.315, p < 0.001). Risk increased with each year of age (O.R. 1.034, p < 0.001). Short women were at decreased risk compared to tall women (O.R. 0.819, p = 0.026). Women of medium height were at decreased risk that was not significant compared to tall women (O.R. 0.929, p = 0.391). HMGA1 SNP rs41269028 had no significant relationship to BC risk (p = 0.602).

|  | 95% L.B. | O.R. | 95% U.B. | p value |
| --- | --- | --- | --- | --- |
| menopause | 3.866 | 4.315 | 4.815 | <0.001 |
| age | 1.030 | 1.034 | 1.038 | <0.001 |
| short | 0.689 | 0.819 | 0.973 | 0.026 |
| medium | 0.788 | 0.929 | 1.095 | 0.391 |
| HMGA1 | 0.908 | 0.980 | 1.058 | 0.602 |

## Discussion

Most of the genetic variation linked to complex phenotypes like height is found in non-coding sections of the genome. 99% of the human genome is non-coding, meaning that the great majority of inherited genetic variation is both uncommon and found there. Identifying the uncommon non-coding variation linked to common features and disorders may help uncover new regulatory gene pathways and significantly advance our knowledge of human biology and disease [4].

HMGA1 SNP rs41269028 is an intron variant in a non-coding region of the genome, chromosome 6. Rare variants such as those of HMGA1 are said to confer most of the heredity for height, about 79% [11]. In other words, in a large group of people 79% of height differences are genetic [12].

HMGA1 is a protein that has been found to play a role in the progression of breast cancer. One study suggests that HMGA1 establishes an autocrine loop in invasive triple-negative breast cancer (TNBC) cells, which mediates the migration, invasion, and metastasis of TNBC cells and predicts the onset of metastasis in these patients [13].

HMGA1 promotes breast cancer angiogenesis by supporting the stability, nuclear localization, and transcriptional activity of FOXM1 [14]. FOXM1 is an oncogenic transcription factor that is greatly upregulated in breast cancer and many other cancers where it promotes tumorigenesis, cancer growth and progression. It is expressed in all subtypes of breast cancer and is the factor most associated with risk of poor patient survival, especially in (TNBC).

Our study has weaknesses. We did not have tumor size, histology, grade, or hormone receptor status. We did not have the recently released UKBB whole genome sequence data for HMGA1 that Hawkes et al used [4, 15]. Instead, we evaluated imputed genotypes from UKB data field 22828 [16]. We found that HMGA1 SNP rs41269028 minor allele T carriers (CT) and homozygotes (TT) were significantly taller, but the effect size was small, 0.59 cm, not the 4.83 cm that Hawkes et al reported [4].

## Conclusion

We conclude that HMGA1 influences height, but we were unable to demonstrate that HMGA1 is related to increased incidence or poor prognosis of tall women with breast cancer. Our finding that tall women have a worse prognosis is important because it could help the oncologist decide, along with other prognostic factors, whether adjuvant therapy is warranted.

## Data Availability

Data sources described in the article are publicly available or can be accessed after approved application to UK Biobank.

https://www.ukbiobank.ac.uk/

## References

1. Gremke N, Griewing S, Kalder M, Kostev K. Positive association between body height and breast cancer prevalence: a retrospective study with 135,741 women in Germany. Breast Cancer Res Treat 2022;196(2):349–354.

2. van den Brandt PA, Ziegler RG, Wang M, et al. Body size and weight change over adulthood and risk of breast cancer by menopausal and hormone receptor status: a pooled analysis of 20 prospective cohort studies. Eur J Epidemiol 2021;36(1):37–55.

3. Unachukwu U, Chada K, D’Armiento J. High mobility group AT-Hook 2 (HMGA2) oncogenicity in mesenchymal and epithelial neoplasia. International journal of molecular sciences 2020;21(9):3151.

4. Hawkes G, Beaumont RN, Li Z, et al. Whole genome association testing in 333,100 individuals across three biobanks identifies rare non-coding single variant and genomic aggregate associations with height. bioRxiv 2023; 10.1101/2023.11.19.566520:2023.11.19.566520.

5. Arthur RS, Wang T, Xue X, et al. Genetic Factors, Adherence to Healthy Lifestyle Behavior, and Risk of Invasive Breast Cancer Among Women in the UK Biobank. J Natl Cancer Inst 2020;112(9):893–901.

6. Mahmood SS, Levy D, Vasan RS, Wang TJ. The Framingham Heart Study and the epidemiology of cardiovascular disease: a historical perspective. Lancet 2014;383(9921):999–1008.

7. Marquez M, Huyvaert M, Perry JR, et al. Low-frequency variants in HMGA1 are not associated with type 2 diabetes risk. Diabetes 2012;61(2):524–30.

8. Chiefari E, Tanyolac S, Paonessa F, et al. Functional variants of the HMGA1 gene and type 2 diabetes mellitus. JAMA 2011;305(9):903–12.

9. Chang CC, Chow CC, Tellier LC, et al. Second-generation PLINK: rising to the challenge of larger and richer datasets. Gigascience 2015;4:7.

10. Sagiv M, Vogelaere PP, Soudry M, Ehrsam R. Role of physical activity training in attenuation of height loss through aging. Gerontology 2000;46(5):266–270.

11. Wainschtein P, Jain D, Zheng Z, et al. Assessing the contribution of rare variants to complex trait heritability from whole-genome sequence data. Nat Genet 2022;54(3):263–273.

12. Geddes L. Genetic study homes in on height’s heritability mystery. Nature 2019;568(7753):444–445.

13. Mendez O, Perez J, Soberino J, et al. Clinical Implications of Extracellular HMGA1 in Breast Cancer. Int J Mol Sci 2019;20(23).

14. Zanin R, Pegoraro S, Ros G, et al. HMGA1 promotes breast cancer angiogenesis supporting the stability, nuclear localization and transcriptional activity of FOXM1. J Exp Clin Cancer Res 2019;38(1):313.

15. Callaway E. World’s biggest set of human genome sequences opens to scientists. Nature 2023;624(7990):16–17.

16. Lehrer S, Rheinstein PH. Association of Kallikrein Related Peptidase 3 (KLK3) gene with dermatophytosis in the UK biobank cohort. Mycoses 2023; 10.1111/myc.13649.

